# Mononeuritis multiplex: an unexpectedly common feature of severe COVID-19

**DOI:** 10.1101/2020.07.19.20149898

**Authors:** Edward Needham, Virginia Newcombe, Andrew Michell, Rachel Thornton, Andrew Grainger, Fahim Anwar, Elizabeth Warburton, David Menon, Monica Trivedi, Stephen Sawcer

**Affiliations:** Cambridge University Hospital NHS Foundation Trust, Hills Road, Cambridge, CB2 0QQ; University Division of Anaesthesia, Department of Medicine, University of Cambridge, CB2 0QQ; University of Cambridge, Department of Clinical Neurosciences, Cambridge Biomedical Campus, Cambridge, CB2 0XY

**Keywords:** COVID-19, neuropathy, nerve injury, mononeuritis multiplex

## Abstract

The prolonged mechanical ventilation required by patients with severe COVID-19 is expected to result in significant Intensive Care Unit – Acquired Weakness (ICUAW) in many of the survivors. However, in our post-COVID-19 follow up clinic we have found that, as well as the anticipated global weakness related to loss of muscle mass, a significant proportion of these patients also have disabling focal neurological deficits relating to an axonal mononeuritis multiplex. Amongst the 69 patients with severe COVID-19 that have been discharged from the intensive care units in our hospital, we have seen 11 individuals (16%) with such neuropathies. In many instances, the multi-focal nature of the weakness in these patients was initially unrecognised as symptoms were wrongly assumed to simply relate to “critical illness neuropathy”. While mononeuropathy is well recognised as an occasional complication of intensive care, our experience suggests that such deficits are common and frequently disabling in patients recovering from COVID-19.

---

The respiratory manifestations of the COVID-19 pandemic have strained health care systems around the world [1]. Thousands of patients have required prolonged periods of mechanical ventilation and many are inevitably emerging with significant “Intensive Care Unit – Acquired Weakness” (ICUAW) [2, 3]. Amongst the COVID-19 survivors attending our ICU follow up clinic we have noticed that, in addition to the anticipated symmetrical weakness related to sarcopenia [4], a significant number of these patients also have marked focal neurological deficits related to superimposed mononeuropathies. Here we present the findings in the first 11 such patients that we have seen (Fig. 1), many of whom are significantly disabled by their neuropathies.

**Figure 1.**
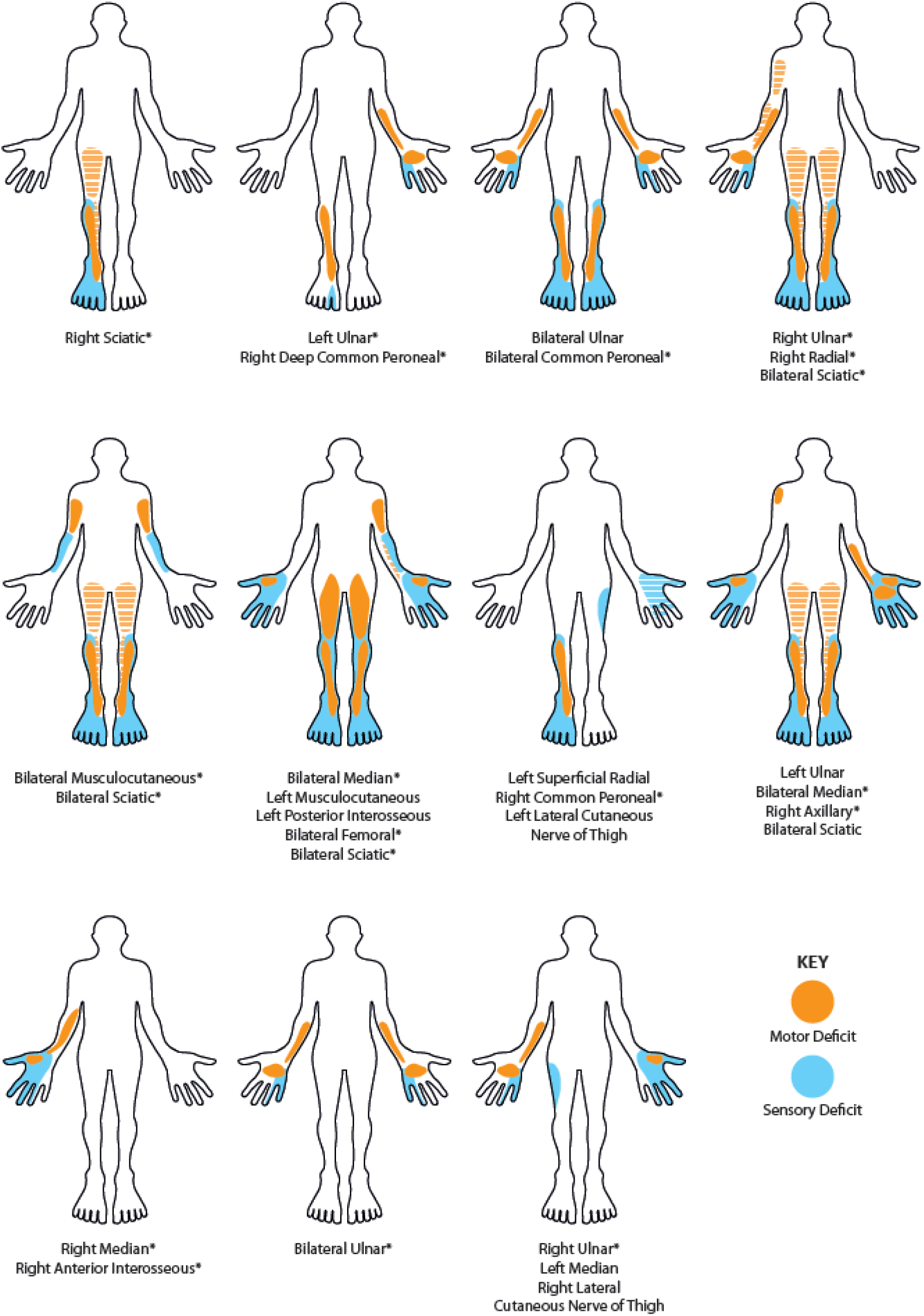
Schematic homuncular illustration of the sensory and motor deficits arising from the multiple mononeuropathies present in eleven patients recovering from severe COVID-19. * denote those neuropathies which were unequivocally confirmed electrophysiologically. Hatched shading indicates posterior muscle groups (hamstrings and triceps). In those neuropathies listed as sciatic there was involvement of both common peroneal and tibial divisions.

All these patients had severe COVID-19 necessitating mechanical ventilation for an average of 31 days (range 16-73), five required periods of proning, and one required extra-corporeal membrane oxygenation (ECMO). Seven of the 11 were men (representative of the known sex difference for severe disease), and the median age was 58 (range 50-77). Four of the patients had type 2 diabetes (none on insulin) and most were overweight, the mean baseline BMI being 26 (range 21-53). The neuropathies were noted following withdrawal of sedation, indicating that these nerve injuries developed during the height of the illness. The clinical deficits have been static or improving when reviewed serially during convalescence, suggesting that the active disease process was limited to the acute phase. In several of the patients, the neuropathies substantially prolonged the need for inpatient rehabilitation and delayed hospital discharge.

All of the patients had evidence of a modest global symmetrical weakness but none showed evidence of significant myopathy on electromyography. In each case, detailed neurophysiological assessment confirmed the presence of additional axonal sensorimotor mononeuropathies, with no evidence of conduction block or other features of neurapraxia. Concomitant generalised neuropathy (typical “critical illness neuropathy” [2]) was only seen in one patient, and this was electrophysiologically mild. Clinically and neurophysiologically, nerve involvement was often patchy with some nerve fascicles more severely affected than others. In those patients with less profound sciatic neuropathies, it was notable that Extensor Hallucis Longus was invariably the most severely affected muscle, out of proportion to the weakness in Tibialis Anterior and Peroneus Longus. Ultrasound scanning of affected nerves was undertaken in three patients and demonstrated diffuse thickening of the affected nerves. The patient with bilateral femoral neuropathies was found to have bilateral psoas haematomata (a complication of ECMO) on MRI, but this finding did not explain the upper limb nor the sciatic neuropathies. Lumbosacral plexus MRI, performed in a second patient with bilateral profound sciatic nerve lesions, was unremarkable: no alternate compressive lesions were identified, and the nerves within the plexus appeared normal. MRI of the brachial plexus in a third patient was also unremarkable. Again, the nerves appeared normal with no evidence of direct injury or avulsion, and no alternative compressive cause identified. As no patient displayed progressive disease, neither nerve biopsy nor lumbar puncture were undertaken. All patients had raised CRP (median 350 [range 268-523]; reference range 0-6), Interleukin 6 (median 28.9pg/ml [range 3.35-400]; reference range 0-2pg/ml), and d-dimer (median 1059ng/ml [range 348-14135]; reference range 0-230ng/ml), but the spectrum of elevation was broad.

To date a total of 102 COVID-19 positive patients have been treated in the ICUs in our hospital, 44 of these have been discharged home, 14 have been transferred to other hospitals for further rehabilitation, 11 are still recovering in our hospital and 33 have died. Not all of the discharged patients have yet been seen for follow up so additional cases of mononeuropathy may still come to light in the coming weeks. Whilst mononeuropathies are, a well-recognised complication of anaesthesia and intensive care^2^, the number of affected patients (at least 16% of those discharged from ICU in our hospital), the number of nerves affected in each patient (an average of just over 3 per patient), and the particular nerves involved (such as the proximal sciatic) are all outside common experience. There have been previous descriptions of bilateral sciatic neuropathies resulting from intensive care, with the patient initially being mistakenly thought to have critical illness neuropathy, but such patients are sufficiently uncommon to warrant individual case reports [5].

## Discussion

This series highlights an important neurological complication of COVID-19, which detrimentally affects the long-term outcomes of patients and markedly influences their rehabilitation needs. Given that this complication is evident in a significant proportion of the patients discharged from the intensive care units of a single hospital (16% in our cohort of treated patients), the rehabilitation burden globally could be substantial. Furthermore, given the high expectation of ICUAW, these focal deficits may go un-noticed.

The underlying aetiology is unclear but the lesions are neurophysiologically axonal, and multifocal, making proning or patient handling related injuries unlikely. Alternative potential explanations would include some form of parainfectious vasculitis as part of the “cytokine storm” seen in COVID [6]. An ischaemic neuropathy resulting from microthrombi in the vasa nervorum [7] might also produce such deficits. The elevations of Interleukin 6 and d-dimer seen in these patients are in keeping with these respective processes, but the broad ranges seen do not clearly support one above the other. However, recent post mortem studies have identified virus-independent immunopathology in cases of fatal COVID-19, and have confirmed that prominent vasculitis occurs in a range of tissues during the illness [8]. In this context, it is reasonable to speculate that the frequency and severity of the mononeuritis multiplex that we have seen in these patients might be less marked in subsequent patients as the standard of care moves to include dexamethasone following the RECOVERY trial [9]. Nerve biopsy may help to further delineate the processes leading to neuropathy, but unless biopsy sites are carefully chosen interpretation of these results is likely to be challenging.

In summary, we have described a mononeuritis multiplex that occurs in a significant fraction of the patients with COVID-19 that have been admitted to the intensive care units of our hospital. While we recognise this is a small number of individuals in total, and that we have no direct comparative group, the clinical features are striking and their implications for rehabilitation profound. We strongly urge detailed neurological assessment of patients with post-COVID-19 ICUAW, especially those with asymmetric weakness, as we suspect that many such patient are likely to have focal deficits resulting from their COVID-19 illness.

## Data Availability

Data is available upon request to the corresponding author.

## Declarations

### Funding

This work was supported by the Cambridge NIHR Biomedical Reseach Centre.

### Conflicts of interest/Competing

None

### Ethics approval

The study was approved by the Cambridge University Hospitals Research and development office.

### Consent to participate

Not applicable (this is not an interventional study)

### Consent for publication

All patients gave fully informed written consent

### Availability of data and material

All available data are included in the paper

### Code availability

Not applicable

### Authors’ contributions

All authors contributes to the clinical care of the patients described and all provided substantive input to the design, drafting and editing of the manuscript

## Acknowledgements

The authors thank Jon Curtis for his help in designing and constructing the figure.

## Notes

### Competing Interest Statement

The authors have declared no competing interest.

### Funding Statement

No funding was received for this work

### Author Declarations

The study was approved by the Cambridge University Hospitals Research and Development office.

